# Knowledge, Attitude and Practices (KAP) towards ketogenic Diet and its risk among Bangladeshi Population: A Cross Sectional Study

**DOI:** 10.1101/2023.08.21.23294352

**Authors:** Arifa Farzana Tanha, Pradip Chandra, Tasnima Haque, Mohammad Mahmudur Rahman

## Abstract

In recent years, the ketogenic diet has become more popular in Bangladesh. Despite the potential negative effects, people are increasingly embracing and adjusting to this diet. Accounting for this manner, the current study addresses a gap that has existed as no research has been done on grown-up adults’ knowledge, attitudes, and practices about the ketogenic diet and its possible risks. To assess their Knowledge, Attitude, and Practice of the Ketogenic Diet and its risks, a descriptive cross-sectional study of the adult Bangladeshi population was carried out. A number of questions were asked to participants regarding their demographics, sources of dietary knowledge, attitudes toward the diet, understanding of the Ketogenic Diet, and views on it. Descriptive statistics were used to analyze the data. Out of 383 participants, women (63.20%) and people under the age of 30 (68.15%) dominated the study. 359 (93.70%) of the participants reported having inadequate knowledge about the ketogenic diet. Around 43.42% of Participants who ever followed KD were assessed to have a healthy BMI. The results revealed a significant association between respondents’ perceptions of KD as a typical weight loss treatment and their knowledge score. Social media was the main source of information for followers (60.53%), while peers were the source of information for 30.26% of respondents. Participants who ever followed the ketogenic diet were less aware of its risks and often considered it safe to follow for a long period believing its benefits outweighed any possible drawbacks. The findings of the current study suggest that Bangladeshis are not well-informed about the therapeutic applications of the ketogenic diet. It appears that pre-diet counselling is essential for everyone to understand every component and proper approach for following this diet.

## 1.0 Introduction

Carbohydrates, in general, are the main source of energy in body systems. When glucose availability in bodily tissues is low, two metabolic processes occur ketogenesis and gluconeogenesis [1, 2]. Gluconeogenesis is the body’s own production of glucose. When glucose availability drops even more, the body’s own production of glucose can’t keep up with the body needs. Hence, ketogenesis starts to make ketone bodies, which are another source of energy. Ketone bodies are used instead of glucose as a main source of energy. Due to low blood glucose feedback during ketogenesis, the stimulus for insulin production is also low, which sharply reduces the stimulus for fat and glucose storage, making it easier for the body to burn fat and store glucose. Several hormonal changes may have a role in the accelerated metabolism of lipids, resulting in the production of fatty acids. Acetoacetate is formed from fatty acids, which is then transformed to beta-hydroxybutyrate and acetone. These are the basic ketone molecules that accumulate in the body when a ketogenic diet is followed for an extended period of time. “Nutritional ketosis” is the name given to this metabolic state. As long as the body is deficient in carbs, the metabolic state stays ketotic. The body’s ketone bodies are readily available for energy synthesis via the heart, muscular tissue, and kidneys the brain. The synthesis of ketones is determined by various variables, including resting basal metabolic rate (BMR), body mass index (BMI), and percentage of body fat. In comparison to glucose, the ketone body makes more adenosine triphosphate, which is often properly referred to as a “super fuel.” One hundred grams of acetoacetate makes 9400 grams of ATP, while 100 grams of beta-hydroxybutyrate make 10,500 grams of ATP. Only 100 grams of glucose makes 8,700 grams of ATP. Even when there is a caloric deficit, the body can continue effective fuel production. In addition, ketone bodies reduce damage caused by free radicals and boost antioxidant capability [3].

The Ketogenic Diet (KD) has a daily nutrition plan that includes meals that are high in fat (accounts for 30–35% of daily caloric need supplying), moderate in protein appropriate (protein minimum of 1 g/kg of protein), low in carbohydrate (carbohydrates offer only 5–10% of total calories, less than 50 g/day) [4, 5]. Dr Russel Wilder first proposed the concept of the Ketogenic Diet in 1921 to manage intractable seizures in paediatric patients [6]. To maintain the metabolic features of ketosis, he proposed the diet with a ratio of ketogenic meals (fats) to anti-ketogenic foods (carbohydrates and protein) greater than 2:1, preferably at least 3:1 [7]. Though, initially the diet included a 4:1 fat-to-carbohydrate and protein ratio [8]. An international study that gathered data from several centres in 17 countries throughout the world, including the Eastern Mediterranean region, has discovered that the overall number of Ketogenic Diet patients in each centre has 71.6, with an annual growth rate of 5.4 patients [4]. However, once antiepileptic medications (AEDs) became accessible, the KD quickly lost favor. It was no longer utilized as a therapeutic option for epilepsy since medicine was simpler to get and consume than the KD [9–11]. Despite this, only around a third of children who use AEDs get epileptic remission [12], hence, during the past 20 years, the KD has come back into use as a different therapy option. Many different types of regimens and diet strategies for bodyweight management have emerged worldwide during the last few years. According to the World Health Organization (WHO), the number of obese patients has tripled since 1975 [4]. KD has gained widespread recognition as one of the most popular strategies for treating obesity since the 1960s [13]. In this manner, the Ketogenic Diet has generated enormous attention among researchers and the general public worldwide [13]. A recent study has found the therapeutic benefits of Ketogenic Diets in various diabetes [14], polycystic ovary syndrome, acne [15, 16], epilepsy [17], cardiovascular, headache [18, 19], metabolic disorders, and the reduction of respiratory in the last century or so [4].

Nevertheless, the ketogenic diet is not regarded as a safe, all-natural, or holistic therapy. Complications are possible, although they are often less and less common than those from medicine or surgery, as with any substantial medical therapy [20]. Additionally, the majority of trials on this diet focused on children who frequently had adverse symptoms such constipation, low-grade acidosis, and hypoglycaemia [21]. Additionally, cholesterol levels may rise by around 30% [20, 22]. The most common short-term side effects of starting the Ketogenic Diet have been dubbed “keto flu“. They include symptoms such as fatigue, headache, dizziness, nausea, vomiting, constipation, and low exercise tolerance [3]. Four weeks following the completion of the ketogenic diet, the researchers identified late consequences including osteopenia, renal stones, cardiomyopathy, secondary hyperornithinaemia, and iron-deficiency anaemia. These complications indicate that the diet is having a negative impact on people, necessitating the need for researchers to conduct more in-depth studies on the diet’s mechanism [23]. Reduced blood insulin levels, increased mitochondriogenesis, and reversal of hepatic insulin resistance in diabetes; increased LDL particles, decreased HMGCoA reductase activity and decreased blood insulin levels in Cardiovascular risk parameters; decreased appetite, decreased lipogenesis, increased lipolysis, the increased metabolic cost of gluconeogenesis, and thermic effect of proteins in weight loss are some of the Ketogenic diet’s outcomes [13].

Despite the pros and cons of the Ketogenic Diet, a significant factor is still missing from the equation: people that follow this diet lack extensive knowledge about the Ketogenic Diet. In other words, a lack of awareness about the ketogenic diet and general practitioner and nutritionist unfamiliarity with the diet can both lead to its misuse [4]. Additionally, improper use of this diet by those who are not good candidates can tarnish its reputation. This, in consequence, will influence the opinions of others who are motivated to utilize it. The ketogenic diet has recently been advocated as a lifestyle choice, although it is highly debatable as to whether doing so is realistic and how long someone can stick with such a diet without suffering negative health or quality of life effects. Hence, the purpose of this study is to determine the level of knowledge, attitude, and practice as well as the risk associated with the ketogenic diet among Bangladeshi residents. It can aid patients in understanding how the ketogenic diet works as a beneficial treatment for a number of medical conditions in addition to weight loss. This study will provide significant empirical data about the practice of the ketogenic diet among adult population in Bangladesh that will help to implement necessary interventions to improve the health and well-being of the people of Bangladesh as no other study has been conducted about knowledge, attitude, and practices parameters regarding the ketogenic diet and its risk among the adult population in Bangladesh.

## 2.0 Methodology

This study was carried out in accordance with the rules established by the Health and Nutrition Organization’s Ethical Review Committee of Institutional Review Board (IRB00013813) (REF# HNO/IRB/2022/NOV/01), dated 5 November 2022. The informed consent form properly detailed the study’s aims, scope, and methods. The participants’ confidentiality and anonymity were strictly maintained.

### 2.1 Study Design, Setting and Participants

The current descriptive cross-sectional study was designed to obtain the information regarding knowledge, attitude and practice towards ketogenic diet and its risk. Data were collected via an online survey through the authors’ social media networks and a face-to-face interview by trained interviewers using a semi-structured questionnaire. The questionnaire consisted of 47 questions, following the study “Knowledge and Perception of Followers of the ketogenic diet among Arab Adults in seventeen Countries” [4]; in accordance with the requirements, the questionnaire was modified. Socio-demographic, knowledge, attitude, practice, and participant views on the ketogenic diet and its risks were the five categories that made up the questionnaire and was divided into sections to guarantee efficient and responsible responses. The questions in the first section (Questions 1–10) covered the sociodemographic information of the participants; the questions in the knowledge, attitude, and practice (Questions 11–21, 22–33, and 34–42) sections covered knowledge, attitude, and practices related to the ketogenic diet and its risks; and the questions in the final section (Questions 43–47), covered the opinions of the participants (who had previously followed the KD). The questionnaire was pre-tested before being distributed to the participants. Some questions received modifications after pre-testing and validation to meet the satisfaction. We removed the required option for questions 36–47 as it would only apply to individuals who had or were on a ketogenic diet. In section four, we also included a question option for participants who had never tried a ketogenic diet, which was number 35. All social media users, who met the criteria were given access to online cross-sectional questionnaires. Volunteers and all authors were involved in the online and offline survey. The survey started on December 1, 2022, and ended on March 1, 2023, among adult people of Bangladesh. Purposive sampling was used to choose a sample of 403 adults from Bangladesh who were ≥18 years old to take part in the study. Except for age, there were no specified requirements for exclusion from this study. Anyone who was at least 18 years old, in good health, and willing to participate in the survey ensued eligible. A total of 400 participants responded; however, 11 participants were excluded from the responded participants due to missing required information. After excluding 9 individuals based on their age during ascending, a final sample of 383 completed questionnaires was chosen for data analysis.

### 2.2 Knowledge, Attitude and Practice (KAP) Questionnaire and their assessment

To assess the respondents’ level of knowledge, 11 questions (Supplementary Table 01) were added to the questionnaire. There were five possible responses for each question: very poor, poor, fair, good, and excellent. Respondents who received a score of 5 out of 5 on a scale of 1 to 5 identified as keeping “excellent knowledge” regarding the ketogenic diet.

### 2.3 Assessments of attitude and practices of respondents

The beliefs and knowledge of respondents about the ketogenic diet among all respondents as well as among respondents who had ever followed the KD were used to measure respondents’ attitudes and practices. Twenty-one questions were used to examine respondents’ attitudes and practices regarding the risks of the ketogenic diet. A total of 20 out of the 21 questions featured a “yes/no” response choice, and in order to collect more detailed responses for 8 questions, there was an option to fill in, “If yes, then please explain“.

### 2.4 Assessments of perceptions

Along with the participants’ responses to certain questions on attitudes and practices, the risk of the ketogenic diet and their perception of the diet’s risks were also assessed. There were 5 questions used to accomplish this.

### 2.5 Handling of variables and processing for analysis

Selected 47 questionnaires were thoroughly reviewed by the authors several times. The ages were divided into two categories: those under 40 and those over 40. Education data were broken down into levels up to HSC/Diploma and levels above HSC/Diploma; occupation data were divided into employed and unemployed (jobless, housewives, and students); and socioeconomic status was divided into Lower and Lower Middle Class and Upper Middle and Upper Class. For analysis, non-numerical data were coded with numbers.

### 2.6 Anthropometric Measurements

Each participant had their body weight and height assessed, and their body mass index (BMI) was determined by dividing their weight in kilograms by their height in meters squared (kg/m2). Healthy BMI was determined to be between 18.5 and 24.9 kg/m2, while unhealthy BMI was determined as <18.5 kg/m2 and >25 Kg/m2.

### 2.7 Statistical Analysis

Excluding noncompliance data, correct data was imported into a database and evaluated using the statistical program SPSS 26 (Statistical Package for Social Science). Descriptive (frequency, percentage, mean, and standard error of mean) analysis was conducted as needed for categorical and quantitative variables. To find the association between socio-demographic characteristics, knowledge and attitude of the all participants and individually those who had practiced ketogenic diet we conducted univariate linear model regression and multivariate logistic regression model. Only KD followers’ individual associations between practice and knowledge were evaluated. A 5% level of significance was utilized. Graph pad prism 8 was used to draw grafts.

## 3.0 Results

### 3.1 Demographical representations

#### 3.1.1 Overall respondents

The 383 research participants were all over the age of 18 years old. The respondents’ average age was determined to be 30.17±0.5998 SEM years. Around 261 respondents, or 68.15% of the sample size, were under 30 years old, which made up the bulk of respondents. About 242 (63.20%) of the responders were female. Higher secondary education (58.70%) was the most common type of education among respondents in terms of educational attainment, followed by postgraduate degrees (18.50%). A large percentage of respondents identified as students (43.90%), while others said they were housewives (23.20%), worked in the private sector (20.10%), or were employed by the government (4.20%). In addition, 23 respondents (6.00%) were jobless, whereas 10 respondents (2.60%) had their own business. 202 (52.70%) of the respondents were now married or had been married in the past. The bulk of those surveyed (60.60%) were from lower-class or lower-middle-class families. Of the total responders, 66 were in the health professions, including 32 doctors and nutritionists and 34 from the categories of nursing, medical assistants, and laboratory workers. All respondents had average body mass indices (BMIs) that were within the healthy range (23.88±0.237 SEM kg/m2). There were some people who were obese (10.20%) and overweight (22.70%) however. A total of 290 respondents (75.70%) stated that they had no chronic illnesses. This study’s full demographic breakdown may be seen in Table 01.

**Table 01:** Demographical distribution of all respondents and respondents who ever followed KD.

| Demography | Group | N (% of total respondents) and Mean±SEM | Following KD Ever |  |  |  |
| --- | --- | --- | --- | --- | --- | --- |
|  |  |  | n and Mean±SEM | % of Total Respondents | % within Demographic Group | % within respondents who have followed KD |
| Respondents |  | 383 (100) | 76 | 19.84 | n/a | 100.00 |
| Age |  | 30.71±0.5998 | 30.07±1.0117 |  |  |  |
| Age Group | Upto 30 Years Old | 261 (68.15) | 48 | 12.53 | 18.39 | 63.16 |
|  | Above 30 Years Old | 122 (31.85) | 28 | 7.31 | 22.95 | 36.84 |
| Gender | Male | 141 (36.80) | 17 | 4.44 | 12.06 | 22.37 |
|  | Female | 242 (63.20) | 59 | 15.40 | 24.38 | 77.63 |
| BMI |  | 23.88±0.237 | 28.80±0.653 |  |  |  |
| BMI Category | Underweight | 40 (10.40) | 6 | 1.57 | 15.00 | 7.89 |
|  | Healthy | 217 (56.70) | 33 | 8.62 | 15.21 | 43.42 |
|  | Overweight | 87 (22.70) | 23 | 6.01 | 26.44 | 30.26 |
|  | Obese | 39 (10.20) | 14 | 3.66 | 35.90 | 18.42 |
| BMI Range | Unhealthy | 166 (43.30) | 43 | 11.23 | 25.90 | 56.58 |
|  | Healthy | 217 (56.70) | 33 | 8.62 | 15.21 | 43.42 |
| Education Category | Primary | 29 (7.60) | 1 | 0.26 | 3.45 | 1.32 |
|  | Secondary | 42 (11.00) | 6 | 1.57 | 14.29 | 7.89 |
|  | Higher Secondary | 225 (58.70) | 49 | 12.79 | 21.78 | 64.47 |
|  | Graduate | 16 (4.20) | 2 | 0.52 | 12.50 | 2.63 |
|  | Postgraduate and above | 71 (18.50) | 18 | 4.70 | 25.35 | 23.68 |
| Education Range | Up to Higher Secondary | 296 (77.30) | 56 | 14.62 | 18.92 | 73.68 |
|  | Above Higher Secondary | 87 (22.70) | 20 | 5.22 | 22.99 | 26.32 |
| Profession Category | Unemployed | 23 (6.00) | 2 | 0.52 | 8.70 | 2.63 |
|  | Student | 168 (43.90) | 27 | 7.05 | 16.07 | 35.53 |
|  | House-wife | 89 (23.20) | 23 | 6.01 | 25.84 | 30.26 |
|  | Govt. Job | 16 (4.20) | 4 | 1.04 | 25.00 | 5.26 |
|  | Private Job | 77 (20.10) | 16 | 4.18 | 20.78 | 21.05 |
|  | Business | 10 (2.60) | 4 | 1.04 | 40.00 | 5.26 |
| Profession Range | Unemployed (including students and House wife) | 280 (73.10) | 52 | 13.58 | 18.57 | 68.42 |
|  | Employed | 103 (26.90) | 24 | 6.27 | 23.30 | 31.58 |
| In Health Profession | Physicians and Nutritionists/Dietitians | 32 (8.40) | 6 | 1.57 | 18.75 | 7.89 |
|  | Other Health Profession (Nurse, Medical Assistant, Laboratory etc.) | 34 (8.90) | 10 | 2.61 | 29.41 | 13.16 |
| Marital Status | Never Married | 181 (47.30) | 35 | 9.14 | 19.34 | 46.05 |
|  | Ever Married | 202 (52.70) | 41 | 10.70 | 20.30 | 53.95 |
| Socioeconomic Status | Lower Class | 27 (7.00) | 3 | 0.78 | 11.11 | 3.95 |
|  | Lower Middle Class | 205 (53.50) | 36 | 9.40 | 17.56 | 47.37 |
|  | Upper Middle Class | 142 (37.10) | 36 | 9.40 | 25.35 | 47.37 |
|  | Upper Class | 9 (2.30) | 1 | 0.26 | 11.11 | 1.32 |
| **Socioeconomic Status Range** | Lower and Lower Middle Class | 232 (60.60) | 39 | 10.18 | 16.81 | 51.32 |
|  | Upper Middle and Upper Class | 151 (39.40) | 37 | 9.66 | 24.50 | 48.68 |
| **Having Long term disease** | Yes | 93 (24.30) | 22 | 5.74 | 23.66 | 28.95 |
|  | No | 290 (75.70) | 51 | 13.32 | 17.59 | 67.11 |
| **Heard about KD** | Yes | 342 (89.30) | 76 | 19.84 | - | 100.00 |
|  | No | 41 (10.70) | 0 | - | - | - |
| **Knowledge Score** |  | 2.15±0.0526 | 2.69±0.0941 |  |  |  |
| **Knowledge Score Category** | Nescience (Score below 3) | 289 (75.50) | 41 | 10.70 | 14.19 | 53.95 |
|  | Average (Score 3 to below 4) | 70 (18.30) | 32 | 8.36 | 45.71 | 42.11 |
|  | Satisfactory (Score 4 to below 5) | 21 (5.50) | 3 | 0.78 | 14.29 | 3.95 |
|  | Excellent (Score 5) | 3 (0.80) | 0 | 0.00 | 0.00 | 0.00 |
| **Knowledge Score Range** | Poor (Score below 4) | 359 (93.70) | 73 | 19.06 | 20.33 | 96.05 |
|  | Good (Score above 4) | 24 (6.30) | 3 | 0.78 | 12.50 | 3.95 |

#### 3.1.2 Ketogenic diet followers

Around 342 (89.30%) respondents had heard about KD before. A total of 76 respondents (19.84%) said that they were either following a ketogenic diet (KD) at the time the study was taken or had previously followed one. These respondents’ average BMI was 28.80±0.653 SEM kg/m2, and their average age was 30.07±1.017 SEM years. This shows that, on average, people who were overweight had at least one experience with the KD. About 48 KD followers, or 63.16% of all KD followers, were under the age of 30, making up 12.53% of all respondents, 18.39% of the same age group, and 63.16% of all KD followers (see Table 01). Around 77.63% of KD supporters were female. About 43.42% of those who followed KD were deemed to have a healthy BMI. The majority of KD supporters (64.47%) had finished their secondary school, with postgraduate responders making up the next-largest section (23.68%). When it comes to occupations, KD followers were more likely to be students (35.53%) and housewives (30.26%) than other types of workers. Ever married respondents (53.95%) had a little higher propensity to adhere to the KD than did those who were never married (46.05%). Socioeconomic classes have no impact on the adoption of this customized diet (Table 01). About 67.11% of KD adherents reported they had no chronic illnesses. It’s interesting to note that 6 doctors and nutritionists were among the 66 health professionals that adhered to the KD.

### 3.2 Knowledge on KD

We used 11 structured questions to assess the knowledge of the responders. On a scale of 1 to 5, the average knowledge score for all participants was 2.15±0.0526 SEM. On the same scale, the mean knowledge score among individuals who have ever followed the KD was 2.69 with a SEM of 0.0941. A score of less than 3 indicated that around 75.50% of respondents lacked proper knowledge of the KD. Only three responders, on the other hand, demonstrated excellent knowledge of the KD (scoring of 5 out of 5). About 359 (93.70%) of the 383 respondents reported inadequate knowledge, which includes both nescience and average knowledge (scoring below 4). However, only 24 respondents (6.30%) showed good knowledge, which includes both of satisfactory and excellent knowledge (scoring of 4 or higher). Three of the 24 respondents with good knowledge have followed the KD at some point in their lives (see Table 1).

#### 3.2.1 KD Knowledge diversity on Age, Gender, Education and Profession

The knowledge of younger respondents (aged 26.33±2.848 SEM years) was superior to that of respondents with nescience knowledge (aged 30.79±0.747 SEM years), those with average knowledge (aged 28.23±1.049 SEM years), and those with satisfactory knowledge (aged 28.62±0.979 SEM years). Nearly half (49.61%) of the responders with nescience knowledge belonged to the under-30 age group (Table 2 lists more knowledge ratings based on age groups). Given that around one-third of respondents were female, Table 2 shows a feminine dominance in the number of females in all knowledge score categories. Similar patterns were seen in the education and career categories, where those with upper secondary education and students dominated. Intriguingly, respondents from the jobless, students, and private profession groups made up an identical amount (n=1) of those with excellent knowledge ratings (Table 2).

**Table 02:** Demographic distribution of knowledge score on KD.

| Parameters and Questions |  | Knowledge Score |  |  |  |  |  |  |  |  |  |  |  |  |  |  |  |
| --- | --- | --- | --- | --- | --- | --- | --- | --- | --- | --- | --- | --- | --- | --- | --- | --- | --- |
|  |  | Nescience (n=289) |  |  |  | Average (n=70) |  |  |  | Satisfactory (n=21) |  |  |  | Excellent (n=3) |  |  |  |
|  |  | A | B | C | D | A | B | C | D | A | B | C | D | A | B | C | D |
| Age |  | 30.79±0.747 |  |  |  | 28.23±1.049 |  |  |  | 28.62±0.979 |  |  |  | 26.33±2.848 |  |  |  |
| Age Group | Up to 30 Years Old (n=261) | 190 | 49.61 | 72.80 | 65.74 | 54 | 14.10 | 20.69 | 77.14 | 15 | 3.92 | 5.75 | 71.43 | 2 | 0.52 | 0.77 | 66.67 |
|  | Above 30 Years Old (n=122) | 99 | 25.85 | 81.15 | 34.26 | 16 | 4.18 | 13.11 | 22.86 | 6 | 1.57 | 4.92 | 28.57 | 1 | 0.26 | 0.82 | 33.33 |
| Gender | Male (n=141) | 115 | 30.03 | 81.56 | 39.79 | 21 | 5.48 | 14.89 | 30.00 | 4 | 1.04 | 2.84 | 19.05 | 1 | 0.26 | 0.71 | 33.33 |
|  | Female (n=242) | 174 | 45.43 | 71.90 | 60.21 | 49 | 12.79 | 20.25 | 70.00 | 17 | 4.44 | 7.02 | 80.95 | 2 | 0.52 | 0.83 | 66.67 |
| BMI |  | 23.74±0.282 |  |  |  | 24.80±0.527 |  |  |  | 23.30±0.569 |  |  |  | 19.58±1.62 |  |  |  |
| BMI Category | Underweight | 34 | 8.88 | 85.00 | 11.76 | 4 | 1.04 | 10.00 | 5.71 | 0 | 0.00 | 0.00 | 0.00 | 2 | 0.52 | 5.00 | 66.67 |
|  | Healthy | 160 | 41.78 | 73.73 | 55.36 | 38 | 9.92 | 17.51 | 54.29 | 18 | 4.70 | 8.29 | 85.71 | 1 | 0.26 | 0.46 | 33.33 |
|  | Overweight | 65 | 16.97 | 74.71 | 22.49 | 20 | 5.22 | 22.99 | 28.57 | 2 | 0.52 | 2.30 | 9.52 | 0 | 0.00 | 0.00 | 0.00 |
|  | Obese | 30 | 7.83 | 76.92 | 10.38 | 8 | 2.09 | 20.51 | 11.43 | 1 | 0.26 | 2.56 | 4.76 | 0 | 0.00 | 0.00 | 0.00 |
| BMI Range | Unhealthy | 129 | 33.68 | 77.71 | 44.64 | 32 | 8.36 | 19.28 | 45.71 | 3 | 0.78 | 1.81 | 14.29 | 2 | 0.52 | 1.20 | 66.67 |
|  | Healthy | 160 | 41.78 | 73.73 | 55.36 | 38 | 9.92 | 17.51 | 54.29 | 18 | 4.70 | 8.29 | 85.71 | 1 | 0.26 | 0.46 | 33.33 |
| Education Category | Primary | 29 | 7.57 | 100.00 | 10.03 | 0 | 0.00 | 0.00 | 0.00 | 0 | 0.00 | 0.00 | 0.00 | 0 | 0.00 | 0.00 | 0.00 |
|  | Secondary | 40 | 10.44 | 95.24 | 13.84 | 2 | 0.52 | 4.76 | 2.86 | 0 | 0.00 | 0.00 | 0.00 | 0 | 0.00 | 0.00 | 0.00 |
|  | Higher Secondary | 159 | 41.51 | 70.67 | 55.02 | 55 | 14.36 | 24.44 | 78.57 | 9 | 2.35 | 4.00 | 42.86 | 2 | 0.52 | 0.89 | 66.67 |
|  | Graduate | 15 | 3.92 | 93.75 | 5.19 | 1 | 0.26 | 6.25 | 1.43 | 0 | 0.00 | 0.00 | 0.00 | 0 | 0.00 | 0.00 | 0.00 |
|  | Postgraduate and above | 46 | 12.01 | 64.79 | 15.92 | 12 | 3.13 | 16.90 | 17.14 | 12 | 3.13 | 16.90 | 57.14 | 1 | 0.26 | 1.41 | 33.33 |
| Education Range | Up to Higher Secondary | 228 | 59.53 | 77.03 | 78.89 | 57 | 14.88 | 19.26 | 81.43 | 9 | 2.35 | 3.04 | 42.86 | 2 | 0.52 | 0.68 | 66.67 |
|  | Above Higher Secondary | 61 | 15.93 | 70.11 | 21.11 | 13 | 3.39 | 14.94 | 18.57 | 12 | 3.13 | 13.79 | 57.14 | 1 | 0.26 | 1.15 | 33.33 |
| Profession Category | Unemployed | 15 | 3.92 | 65.22 | 5.19 | 4 | 1.04 | 17.39 | 5.71 | 3 | 0.78 | 13.04 | 14.29 | 1 | 0.26 | 4.35 | 33.33 |
|  | Student | 119 | 31.07 | 70.83 | 41.18 | 40 | 10.44 | 23.81 | 57.14 | 8 | 2.09 | 4.76 | 38.10 | 1 | 0.26 | 0.60 | 33.33 |
|  | House-wife | 80 | 20.89 | 89.89 | 27.68 | 9 | 2.35 | 10.11 | 12.86 | 0 | 0.00 | 0.00 | 0.00 | 0 | 0.00 | 0.00 | 0.00 |
|  | Govt. Job | 9 | 2.35 | 56.25 | 3.11 | 4 | 1.04 | 25.00 | 5.71 | 3 | 0.78 | 18.75 | 14.29 | 0 | 0.00 | 0.00 | 0.00 |
|  | Private Job | 57 | 14.88 | 74.03 | 19.72 | 12 | 3.13 | 15.58 | 17.14 | 7 | 1.83 | 9.09 | 33.33 | 1 | 0.26 | 1.30 | 33.33 |
|  | Business | 9 | 2.35 | 90.00 | 3.11 | 1 | 0.26 | 10.00 | 1.43 | 0 | 0.00 | 0.00 | 0.00 | 0 | 0.00 | 0.00 | 0.00 |
| Profession Range | Unemployed (including student and House wife) | 214 | 55.87 | 76.43 | 74.05 | 53 | 13.84 | 18.93 | 75.71 | 11 | 2.87 | 3.93 | 52.38 | 2 | 0.52 | 0.71 | 66.67 |
|  | Employed | 75 | 19.58 | 72.82 | 25.95 | 17 | 4.44 | 16.50 | 24.29 | 10 | 2.61 | 9.71 | 47.62 | 1 | 0.26 | 0.97 | 33.33 |
| In Health Profession | Physicians and Nutritionists/Dietitians | 14 | 3.66 | 43.75 | 4.84 | 7 | 1.83 | 21.88 | 10.00 | 10 | 2.61 | 31.25 | 47.62 | 1 | 0.26 | 3.13 | 33.33 |
|  | Other Health Profession (Nurse, Medical Assistant, Laboratory etc.) | 22 | 5.74 | 64.71 | 7.61 | 10 | 2.61 | 29.41 | 14.29 | 2 | 0.52 | 5.88 | 9.52 | 0 | 0.00 | 0.00 | 0.00 |
| Marital Status | Never Married | 143 | 37.34 | 79.01 | 49.48 | 47 | 12.27 | 25.97 | 67.14 | 9 | 2.35 | 4.97 | 42.86 | 3 | 0.78 | 1.66 | 100.00 |
|  | Ever Married | 146 | 38.12 | 72.28 | 50.52 | 23 | 6.01 | 11.39 | 32.86 | 12 | 3.13 | 5.94 | 57.14 | 0 | 0.00 | 0.00 | 0.00 |
| **Socioeconomic Status Category** | Lower Class | 25 | 6.53 | 92.59 | 8.65 | 1 | 0.26 | 3.70 | 1.43 | 1 | 0.26 | 3.70 | 4.76 | 0 | 0.00 | 0.00 | 0.00 |
|  | Lower Middle Class | 163 | 42.56 | 79.51 | 56.40 | 37 | 9.66 | 18.05 | 52.86 | 5 | 1.31 | 2.44 | 23.81 | 0 | 0.00 | 0.00 | 0.00 |
|  | Upper Middle Class | 95 | 24.80 | 66.90 | 32.87 | 31 | 8.09 | 21.83 | 44.29 | 13 | 3.39 | 9.15 | 61.90 | 3 | 0.78 | 2.11 | 100.00 |
|  | Upper Class | 6 | 1.57 | 66.67 | 2.08 | 1 | 0.26 | 11.11 | 1.43 | 2 | 0.52 | 22.22 | 9.52 | 0 | 0.00 | 0.00 | 0.00 |
| **Socioeconomic Status Range** | Lower and Lower Middle Class | 188 | 49.09 | 81.03 | 65.05 | 38 | 9.92 | 16.38 | 54.29 | 6 | 1.57 | 2.59 | 28.57 | 0 | 0.00 | 0.00 | 0.00 |
|  | Upper Middle and Upper Class | 101 | 26.37 | 66.89 | 34.95 | 32 | 8.36 | 21.19 | 45.71 | 15 | 3.92 | 9.93 | 71.43 | 3 | 0.78 | 1.99 | 100.00 |
| **Having Long term disease** | Yes | 75 | 19.58 | 80.65 | 25.95 | 15 | 3.92 | 16.13 | 21.43 | 2 | 0.52 | 2.15 | 9.52 | 1 | 0.26 | 1.08 | 33.33 |
|  | No | 214 | 55.87 | 73.79 | 74.05 | 55 | 14.36 | 18.97 | 78.57 | 19 | 4.96 | 6.55 | 90.48 | 2 | 0.52 | 0.69 | 66.67 |
| **What types of nutrient deficiencies may happen while maintain KD** | Only Macronutrients | 48 | 12.53 | - | 16.61 | 13 | 3.39 | - | 18.57 | 9 | 2.35 | - | 42.86 | 1 | 0.26 | - | 33.33 |
|  | Only Micronutrients | 29 | 7.57 | - | 10.03 | 15 | 3.92 | - | 21.43 | 11 | 2.87 | - | 52.38 | 3 | 0.78 | - | 100.00 |
|  | Both Macro and Micronutrients | 19 | 4.96 | - | 6.57 | 5 | 1.31 | - | 7.14 | 5 | 1.31 | - | 23.81 | 1 | 0.26 | - | 33.33 |
| **What diseases can be treated by KD** | Epilepsy | 4 | 1.04 | - | 1.38 | 3 | 0.78 | - | 4.29 | 6 | 1.57 | - | 28.57 | 0 | 0.00 | - | 0.00 |
|  | Diabetes | 39 | 10.18 | - | 13.49 | 8 | 2.09 | - | 11.43 | 4 | 1.04 | - | 19.05 | 3 | 0.78 | - | 100.00 |
|  | Heart Disease and Hypertension | 17 | 4.44 | - | 5.88 | 4 | 1.04 | - | 5.71 | 0 | 0.00 | - | 0.00 | 3 | 0.78 | - | 100.00 |
|  | Overweight and Obesity | 13 | 3.39 | - | 4.50 | 4 | 1.04 | - | 5.71 | 0 | 0.00 | - | 0.00 | 0 | 0.00 | - | 0.00 |
|  | Cancer | 2 | 0.52 | - | 0.69 | 0 | 0.00 | - | 0.00 | 1 | 0.26 | - | 4.76 | 0 | 0.00 | - | 0.00 |
|  | Asthma | 1 | 0.26 | - | 0.35 | 0 | 0.00 | - | 0.00 | 0 | 0.00 | - | 0.00 | 0 | 0.00 | - | 0.00 |
|  | Polycystic Ovary Syndrome | 1 | 0.26 | - | 0.35 | 1 | 0.26 | - | 1.43 | 2 | 0.52 | - | 9.52 | 0 | 0.00 | - | 0.00 |
|  | Autism | 1 | 0.26 | - | 0.35 | 0 | 0.00 | - | 0.00 | 3 | 0.78 | - | 14.29 | 0 | 0.00 | - | 0.00 |
|  | Allergy | 1 | 0.26 | - | 0.35 | 0 | 0.00 | - | 0.00 | 0 | 0.00 | - | 0.00 | 0 | 0.00 | - | 0.00 |
|  | Acidity | 1 | 0.26 | - | 0.35 | 0 | 0.00 | - | 0.00 | 0 | 0.00 | - | 0.00 | 0 | 0.00 | - | 0.00 |
|  | Fatty Liver Disease | 1 | 0.26 | - | 0.35 | 0 | 0.00 | - | 0.00 | 0 | 0.00 | - | 0.00 | 0 | 0.00 | - | 0.00 |
|  | Kidney Diseases | 0 | 0.00 | - | 0.00 | 2 | 0.52 | - | 2.86 | 1 | 0.26 | - | 4.76 | 2 | 0.52 | - | 66.67 |
|  | Constipation | 0 | 0.00 | - | 0.00 | 1 | 0.26 | - | 1.43 | 0 | 0.00 | - | 0.00 | 0 | 0.00 | - | 0.00 |
|  | Alzheimer's Disease | 0 | 0.00 | - | 0.00 | 0 | 0.00 | - | 0.00 | 2 | 0.52 | - | 9.52 | 0 | 0.00 | - | 0.00 |
|  | Parkinson's Diseases | 0 | 0.00 | - | 0.00 | 0 | 0.00 | - | 0.00 | 2 | 0.52 | - | 9.52 | 0 | 0.00 | - | 0.00 |
|  | Neurological Disorder | 0 | 0.00 | - | 0.00 | 0 | 0.00 | - | 0.00 | 1 | 0.26 | - | 4.76 | 0 | 0.00 | - | 0.00 |
|  | Metabolic Disorder | 0 | 0.00 | - | 0.00 | 0 | 0.00 | - | 0.00 | 2 | 0.52 | - | 9.52 | 1 | 0.26 | - | 33.33 |
| **Do you think KD should be followed by the supervision of a nutritionist?** | Yes | 130 | 33.68 | 77.71 | 44.64 | 32 | 8.36 | 19.28 | 45.71 | 3 | 0.78 | 1.81 | 14.29 | 2 | 0.52 | 1.20 | 66.67 |
|  | No | 159 | 41.78 | 73.73 | 55.36 | 38 | 9.92 | 17.51 | 54.29 | 18 | 4.70 | 8.29 | 85.71 | 1 | 0.26 | 0.46 | 33.33 |

A = n and Mean±SEM; B = % of Total Respondents; C = % within Demographic Group; D = % within knowledge score group

#### 3.2.2 KD Knowledge in Health Profession

A total of 66 respondents from the health professions participated in the survey, and 36 of them demonstrated nescience knowledge—14 of them were doctors and nutritionists and 22 were from other health professions. Surprisingly, Table 2’s results for the physicians and nutritionists respondents only indicate one responder who had excellent knowledge.

#### 3.2.3 KD Knowledge diversity on Marital and Socioeconomic Status

The respondents with nescience knowledge were virtually equally split by marital status, with 49.48% never married and 50.52% ever married. However, as seen in Table 2, none of the respondents with excellent knowledge had ever been married. The majority of the 163 respondents from the lower middle class in the Socioeconomic Status Category showed nescience knowledge. In contrast, the group of upper middle-class respondents (n=3) included the respondents with excellent knowledge. The lowest and lower-middle classes of the socioeconomic class made up around 65.05% of the group with nescience knowledge. Table 2 has further information.

#### 3.2.4 KD Knowledge diversity on Diseases and Health Conditions

The average BMI of each knowledge group—nescience (23.74±0.282 SEM BMI), average (24.80±0.527 SEM BMI), satisfactory (23.30±0.569 SEM BMI), and excellent (19.58±1.62 SEM BMI)—fell within the healthy BMI range. About 160 people, or 55.36% of those who responded to the nescience knowledge survey, had a healthy BMI. Around 34 were categorized as underweight, 65 as overweight, and 30 as obese among the 129 remaining (44.64%) nescience knowledge responders. Refer to Table 2 for further information on the connection between knowledge groups and BMI categories. According to Table 2, about 25.95% of those who claimed having nescience knowledge, 21.43% of those who reported having average knowledge, 9.52% of those who reported having satisfactory knowledge, and 33.33% of those who reported having excellent knowledge reported having long-term disorders. As part of our study, we asked the respondents about the kinds of nutritional deficits that could happen when maintaining a KD. Multiple choices were available to the responders. Only 19 people (6.57%) in the nescience knowledge group gave answers that addressed both macro and micronutrients. In a similar vein, responses were provided on both macro and micronutrients by 5 respondents (7.14%), 5 respondents (23.81%), and 1 respondent (33.33%) from the groups with average, satisfactory, and excellent knowledge, respectively. In Table 2 and Figure 1a, more information is provided.

**Figure 1:**
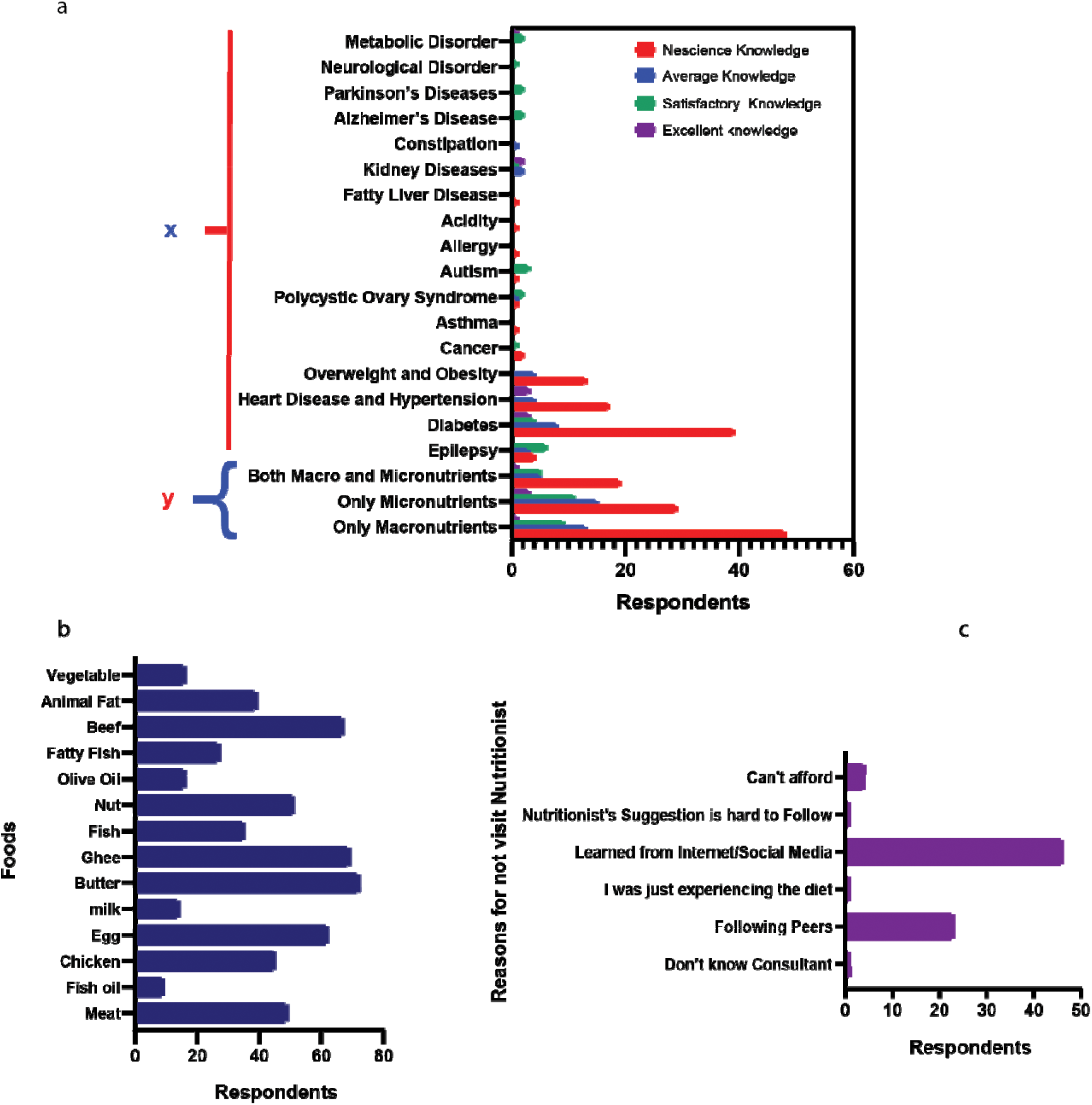
a) responses of participants on knowledge on x = which types of nutrient deficiency can be observed during KD, y = which diseases can be treated with KD; b) what types of foods respondents took during KD; c) Why respondents did not go for nutritionist’s suggestions

Additionally, we asked what ailments a KD may address. Multiple responses were available for respondents to choose from. Thirteen respondents selected epilepsy, 54 selected diabetes, 24 selected heart disease and hypertension, seventeen mentioned overweight and obesity, five selected kidney diseases, four mentioned polycystic ovary syndrome, four mentioned autism, three mentioned cancer, three mentioned metabolic disorders, and two mentioned Parkinson’s disease and neurological disorders. Additionally, one responder each stated neurological condition, acidity, fatty liver disease, acidity, asthma, and allergies. Table 2 and Figure 1a show how the scores for knowledge and illnesses are distributed.

### 3.3 Association of Knowledge Score with Demography, Health and Disease Conditions

The findings of both univariate and multivariate analyses are shown in Table 3, which looks at the association between Knowledge Score and different demographic characteristics, health problems, and disease conditions among all respondents, including those who have ever followed the KD.

**Table 03:** Association of Demographic Parameters with Knowledge among Entire Participants and with respondents who ever followed KD.

|  | Knowledge Score vs Demography in All Respondents |  |  |  |  |  |  |  | Knowledge Score vs Demography in Respondents who have followed KD |  |  |  |
| --- | --- | --- | --- | --- | --- | --- | --- | --- | --- | --- | --- | --- |
| Parameter | OR | 95% Confidence Interval |  | Sig. | AOR | 95% Confidence Interval |  | Sig. | OR | 95% Confidence Interval |  | Sig. |
|  |  | Lower Bound | Upper Bound |  |  | Lower Bound | Upper Bound |  |  | Lower Bound | Upper Bound |  |
| Age Group<br>(Up to 30 years old =0), Above 30 years old =1 | -0.003 | -0.068 | 0.061 | 0.920 | 0.713 | 0.199 | 2.560 | 0.604 | 0.076 | -0.057 | 0.209 | 0.259 |
| Gender<br>(male = 0, female =1) | 0.071 | 0.019 | 0.123 | 0.008 | 5.271 | 1.505 | 18.457 | 0.009 | 0.152 | 0.016 | 0.288 | 0.029 |
| BMI Range<br>(unhealthy =0, healthy = 1) | 0.066 | 0.020 | 0.113 | 0.006 | 6.582 | 1.844 | 23.491 | 0.004 | 0.119 | 0.024 | 0.214 | 0.015 |
| Education Range<br>(Up to Higher Secondary = 0, Above Higher Secondary = 1) | 0.095 | 0.030 | 0.160 | 0.004 | 4.535 | 1.538 | 13.367 | 0.006 | 0.081 | -0.042 | 0.205 | 0.192 |
| Profession<br>(Unemployed = 0, Employed = 1) | 0.000 | -0.068 | 0.067 | 0.992 | 1.754 | 0.484 | 6.356 | 0.392 | -0.026 | -0.159 | 0.107 | 0.695 |
| Health Profession<br>(Yes = 1, No = 0) | 0.144 | 0.078 | 0.209 | 0.000 | 6.681 | 2.348 | 19.012 | 0.000 | 0.112 | -0.028 | 0.252 | 0.115 |
| Marital Status<br>(never married =0, ever married =1) | -0.001 | -0.059 | 0.058 | 0.981 | 1.102 | 0.386 | 3.145 | 0.856 | -0.101 | -0.235 | 0.033 | 0.136 |
| Socioeconomic Range<br>(Lower and Lower Middle = 0, Upper Middle and Upper = 1) | 0.070 | 0.022 | 0.118 | 0.004 | 5.018 | 1.696 | 14.845 | 0.004 | 0.058 | -0.034 | 0.149 | 0.211 |
| Having Diseases<br>(Yes =1, No = 0) | -0.042 | -0.098 | 0.014 | 0.142 | 0.387 | 0.090 | 1.672 | 0.204 | 0.003 | -0.099 | 0.106 | 0.949 |

Poor Knowledge Score is Reference. (Good Knowledge = 1, Poor Knowledge =0)

The following variables across all respondents shown significant associations with the knowledge score:

1. Gender: The odds ratio (OR) was 0.071, and the 95% confidence interval (CI) ranged from (0.019 to 0.123). The p-value for the analysis was 0.008. The connection remained significant after correcting for other covariates (AOR), with an AOR of 5.271, 95% CI (1.505 - 18.457), and a p-value of 0.009.
2. BMI Range: With a p-value of 0.006, the OR was 0.066, 95% CI (0.020 - 0.113), and the results. The AOR was 6.582, 95% CI (1.844 - 23.491), and p-value was 0.004 after controlling for other variables.
3. The OR was 0.095, with a 95% confidence interval (0.030-0.160), and a p-value of 0.004 for the education range. The AOR was 4.535, 95% CI (1.538 - 13.367), with a p-value of 0.006 after correcting for other variables.
4. Health Profession: With a p-value of 0.000, the OR for this group was 0.144, 95% CI (0.078-0.209). The adjusted odds ratio (AOR) was 6.681, with a 95% confidence interval (CI) of (2.348 to 19.012) and a p-value of 0.000.
5. The OR was 0.070, 95% CI (0.022-0.118), with a p-value of 0.004 for the socioeconomic range. The AOR was 5.018, 95% CI (1.696 - 14.845), and p-value was 0.004 after correcting for other variables.

Based on these findings, it can be concluded that among all respondents, the knowledge score was strongly influenced by gender, BMI range, educational level, health profession, and socioeconomic position. However, only two variables indicated significant relationships with the knowledge score among respondents who had ever used the KD:

1. Gender: A p-value of 0.029, an odds ratio (OR) of 0.152, and a 95% confidence interval (CI) of (0.016 to 0.288), were the results. This shows that among respondents who have ever followed the KD, gender has a meaningful relationship with knowledge score.
2. BMI Range: With a p-value of 0.015, the OR was 0.119, 95% CI (0.024-0.214). This suggests that among respondents who have ever followed the KD, BMI range likewise has a strong correlation with the knowledge score.

These results suggest that among respondents who have ever followed the KD, the only variables that exhibited significant associations with the knowledge score were gender and BMI range.

### 3.4 Association of Knowledge Score with Attitude towards KD

We wanted to find out if respondents’ perceptions of KD and its implications on health were connected to their knowledge of the condition. We probed them with 12 questions to gauge their opinions. We used both univariate and multivariate regression analyses to comprehend the connection between KD knowledge score and attitude among all respondents as well as among respondents who had ever followed KD. The fact that respondents thought the KD diet would put people at risk for dietary deficiencies was one interesting conclusion. A link between knowledge score and odds ratio (OR) was found to be significant (OR: 0.080, 95% CI: 0.023-0.137, p-value: 0.006). The data, however, demonstrated the opposite trend after correcting for other variables (Adjusted Odds Ratio - AOR: 0.427, 95% CI: 0.107 - 1.702, p-value: 0.228). The results show a complicated and intriguing link between respondents’ views regarding KD and their knowledge of it, with the adjusted odds pointing in a different direction than the first univariate study. Regarding the following topics, similar correlations between respondents’ beliefs and their knowledge score were discovered:

1. The knowledge score of the respondents significantly correlated with the respondents’ perception that the advantages of KD outweigh the risks (OR: -0.063, 95% CI: -0.116 - - 0.010, p-value: 0.019). The link did not remain significant after controlling for other variables, with an AOR of 5.837, 95% CI: 0.779 - 43.709, and a p-value of 0.086.
2. There was a significant link between respondents’ knowledge score and their view that one should discontinue the KD after they lose weight (OR: 0.053, 95% CI: 0.007-0.099, p-value: 0.025). With an AOR of 0.252, 95% CI: 0.056 - 1.143, and a p-value of 0.074, the relationship did not exhibit a tendency towards significance after controlling for other variables.

Furthermore, it was discovered that respondents’ perception of KD as a typical weight reduction strategy was substantially correlated with their knowledge score (OR: -0.051, 95% CI: -0.107 - 0.004, p-value: 0.071). The association remained significant after controlling for other variables, with an AOR of 7.699, 95% CI: 1.157 - 51.236, and a p-value of 0.035. Additionally, respondents’ knowledge score was substantially correlated with their view that KD may treat illnesses (OR: 0.181, 95% CI: 0.130 - 0.233, p-value: 0.000). The connection remained highly significant after accounting for other covariates, with an AOR of 0.033, 95% CI: 0.009 - 0.119, and a p-value of 0.000.The correlation between respondents’ beliefs and knowledge scores is highlighted by these data, highlighting the significance of attitudes and perceptions in relation to KD.

The knowledge scores of respondents who have ever used the ketogenic diet (KD) are significantly correlated with their attitudes toward the notion that their weight will remain the same when they stop using the KD (OR: 0.200, 95% CI (0.055 - 0.344), p-value: 0.008) and the notion that the KD can be used to treat diseases (OR: 0.152, 95% CI (0.039 - 0.265), p-value: 0.009). In the full set of respondents and those who had ever followed the KD; however, no statistically significant relationships were discovered between their knowledge scores and any other attitudes (Table 4).

**Table 04:**
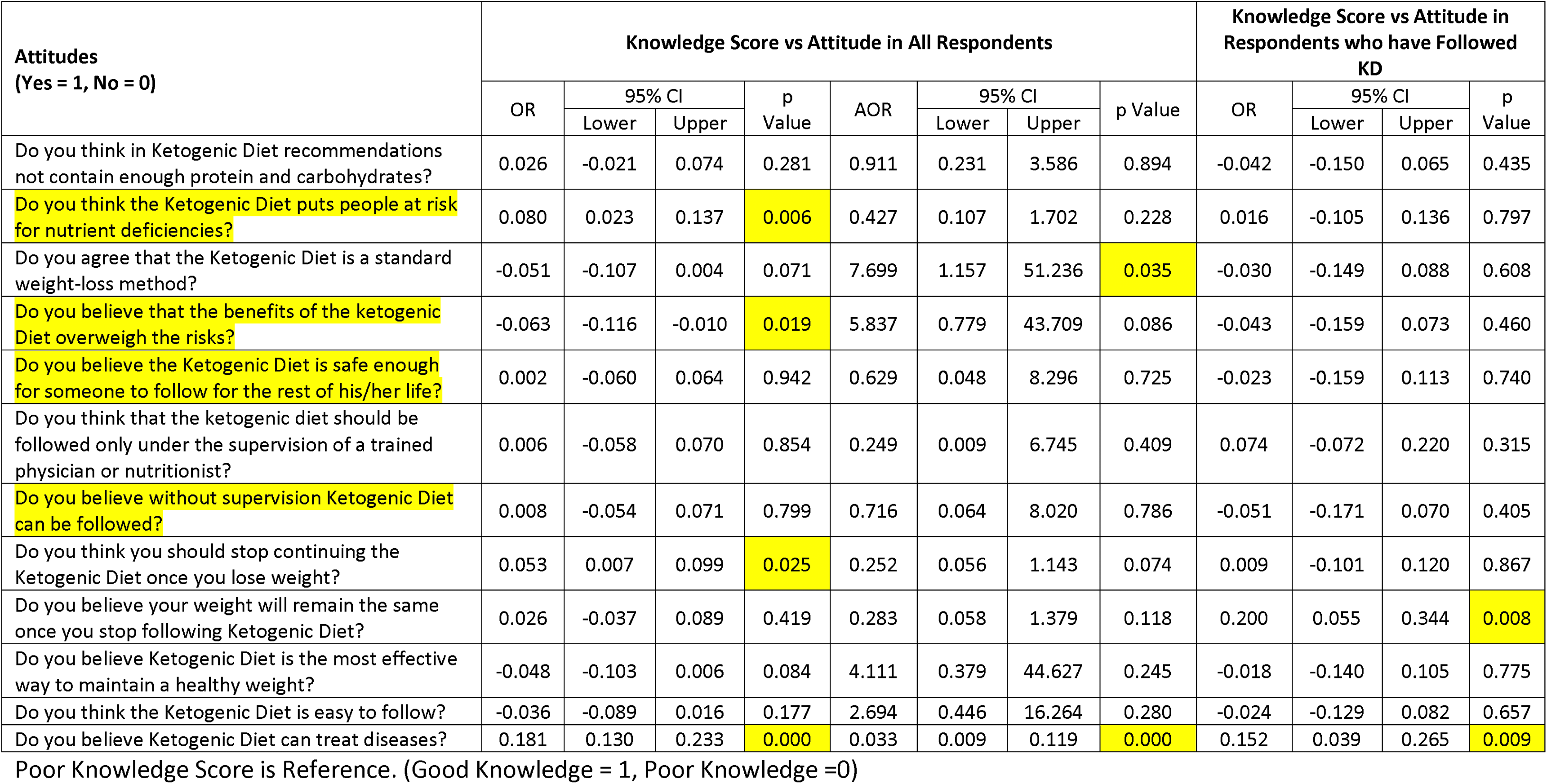
Association of Attitude with Knowledge among Entire Participants and with respondents who ever followed KD.

### 3.5 Association of Knowledge Score with Practices on KD

As expected, not any significant connections between the respondents’ practices and their knowledge were discovered. There were seven questions in all, and none of the individual practices revealed any kind of statistically significant link (Table 5).

**Table 05:**
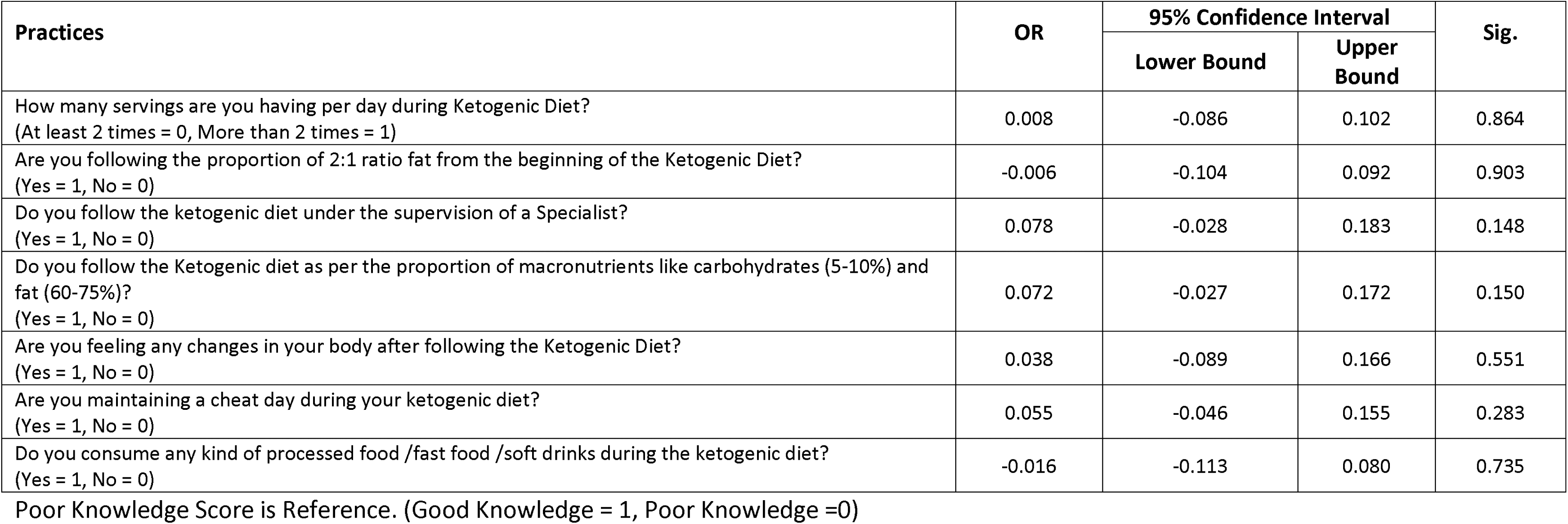
Association of Practices with Knowledge among respondents who ever followed KD.

### 3.6 Perspectives of Respondents on KD

Additional questions were asked of respondents who had ever used the ketogenic diet (KD) in order to learn more about their viewpoints, eating habits, length of time spent adhering to the KD, experiences during and after the KD, and, if appropriate, the reasons for stopping the KD. Multiple responses were available for certain questions. The majority of responders focused on eating butter, ghee, meat, and eggs as their main meal selections during the KD (Figure 1b). Furthermore, they consumed meat, fish, chicken, and animal fats as part of their diet. The dietary choice that people liked the least was fish oil. A total of 43 (60.53%) respondents were found to have learnt about and adopted KD recommendations from the internet and social media, whereas 23 (30.26%) respondents had sought advice from their peers. Three respondents said they couldn’t afford the services of a nutritionist or expert, and just one respondent said they sought guidance from a nutritionist (Figure 1c). The majority (18) of the respondents followed the KD for three months (Figure 2a). About 32 respondents (42.11%) stated that they actually lost weight, but around a third (23, 30.26%) said they felt unwell, fainted, and dizzy throughout the KD (Figure 2b). Nine (11.84%) of the 76 responders who had previously followed the KD discontinued after attaining their goal weight loss, whereas about 18 (23.68%) continued the diet. However, owing to health-related reasons, almost 20 (26.32%) responders stopped the KD. Lack of ability (8, 10.53%), hair loss (5, 6.58%), and awareness of its negative consequences (3, 3.95%) were other reasons for quitting the KD. Figure 2c presents more justifications.

**Figure 2:**
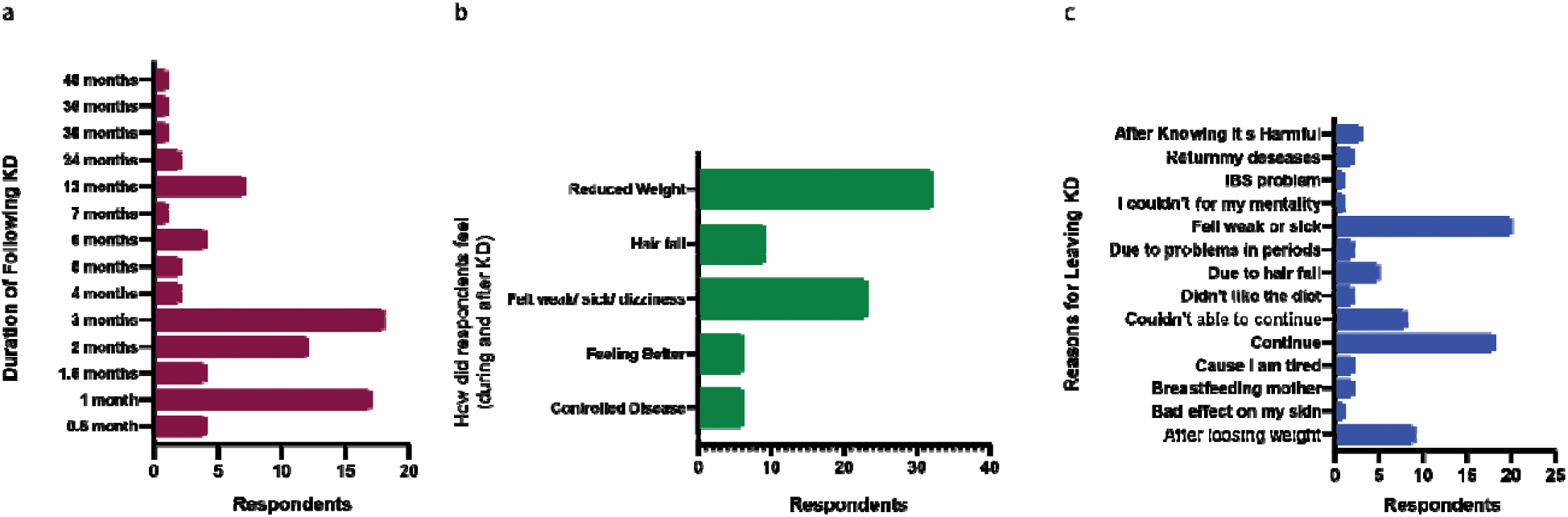
a) Duration of KD following; b) How did respondents feel during KD; c) Why respondent left KD

## 4.0 Discussion

The ketogenic diet may be helpful in treating a number of medical disorders [13], especially in children with difficult-to-treat epilepsy [24, 25], although general practitioners most frequently associate it with patient weight reduction. Indeed, ketogenic diets (KDs) have demonstrated success, particularly in the short to medium term, in addressing issues like obesity and overweight [26], metabolic disorders like hyperlipidemia, cardiovascular risks, and some neurological conditions [27–31]. However, the KD raises serious concerns among the medical profession and is not advised for wider adoption by the public [32]. Many of these concerns are probably the result of incomplete knowledge of the physiological processes behind therapeutic ketogenic diets.

This study set out to assess how well-informed, conscious, and aware the Bangladeshi populace was of KD. Women made up the bulk of the study’s participants (n=242, 63.20%) due to their focus on weight control and ambitions for the ideal body form. This pattern is consistent with prior studies that examined knowledge and beliefs of KD and mostly included female respondents [33–36]. Notably, the survey found that a large percentage of participants (89.30%) were previously familiar with KD, which is comparable to the results of a research performed in Pakistan (73.2%) (Butt et al., 2020). This result highlights how commonplace the ketogenic diet is in Bangladeshi society.

People typically choose diets to lose extra weight, frequently placing this objective ahead of any adverse effects or health consequences. Referring to Table 4, we found that participants in our study saw the ketogenic diet as a tried-and-true strategy for losing weight. Earlier cross-sectional studies carried out in Pakistan and Palestine produced findings that were comparable [29, 34, 37]. These studies support our own findings and emphasize that losing weight is the main goal of following a ketogenic diet. Notably, a large proportion of individuals who followed the Ketogenic Diet found it to be favorable for controlling chronic disorders such as diabetes and cardiovascular diseases (as seen in Table 4 and Figure 1). This demonstrates the Ketogenic Diet’s wide acceptance and perceived effectiveness in promoting quick weight reduction. Furthermore, compared to low-calorie, low-fat diets, a low-carb ketogenic diet had larger weight reduction advantages, according to a study including obese people [38].

Studies carried out in many countries have shown that different young people follow different low-carb diets [34]. For instance, a Brazilian research [39] found that 25% of students consumed low-carbohydrate foods while having an average BMI that was within normal range. All individuals in our research had average BMIs that were within the normal range (23.88±0.237). However, the average BMI among respondents who had ever used the ketogenic diet (about 20% of all respondents) was 28.80±0.653. This conflicting outcome is probably a result of the respondents’ varied backgrounds, including their ages, levels of education, socioeconomic position, and other characteristics. These results highlight the value of promoting healthy eating, realistic weight goals, and developing a good self-image. We found that the research participants didn’t have a thorough understanding of the ketogenic diet. While a sizable portion of respondents thought the diet may treat diabetes, the majority were ignorant of the diet’s ability to treat epilepsy. It’s interesting to note that people who had used the ketogenic diet showed little understanding of its dangers, often believing that its advantages outweighed any potential negatives and that it was safe to take for a long time. As a result, there is a definite need for increased informational and public awareness of the potential dangers of the ketogenic diet.

Internet and social media were the most popular places to learn about the Ketogenic Diet, followed by peer pressure. In research carried out in the UK, social media was notably listed as the least frequent source of nutritional information [36]. In contrast, studies conducted in the UK [36] and Palestine [34] found that the majority of participants had attended nutrition-related courses while pursuing their higher education. In Bangladesh, on the other hand, such instruction is exclusively provided in university settings to students pursuing courses in nutrition and food. NGOs do spread some nutrition information, but it is clear that their efforts fall short. This implies that dietary information should be included in educational programs at all levels of schooling. This could make it easier for people to distinguish between good and harmful meals and motivate them to seek the advice of nutritionists or diet counselors before starting a therapeutic diet. According to Table 2, 43.60% of respondents believed that following the Ketogenic Diet should be done under a nutritionist’s supervision. Interesting results from a UK research [36] showed that professional supervision was required for people following the ketogenic diet, with over 60% of respondents expressing this opinion. The causes of these variations are clear and have already been covered.

While many students believed the ketogenic diet to be difficult to follow or maintain, there ought to be a wider range of views among them on its compliance and simplicity. Similar findings were made in other research [36], when participants mentioned typical adverse effects of the ketogenic diet as hair loss, weakness, dizziness, and more. Due to calorie limits and lower energy levels, these symptoms are frequently linked to different diet regimens. The Ketogenic Diet may raise the risk of heart disease, low blood pressure, kidney stones, constipation, nutritional shortages, and other health problems. Strict eating plans like the ketogenic diet may also contribute to eating disorders or social isolation. It’s significant to mention that anyone with disorders affecting their thyroid, gallbladder, liver, or pancreas should avoid the ketogenic diet. The “keto flu,” which is marked by symptoms including an upset stomach, dizziness, low energy, and mood changes while the body adjusts to ketosis, may be experienced by those who are new to the ketogenic diet [40].

There were 66 members of the healthcare industry among the attendees. 32 of them were doctors or dietitians, while the rest 34 were employed in a range of other healthcare positions, including nursing, medical assistance, and laboratory work (see Table 1). Six of the healthcare participants in the group were doctors or nutritionists, and 10 came from other healthcare specialties and had prior experience with the ketogenic diet (KD). Surprisingly, out of the 32 doctors and nutritionists, only 1 had great understanding of KD, and 14 (representing 43.75 percent of the doctors/nutritionists) had insufficient knowledge (see Table 2). This findings points to a general lack of thorough dietary knowledge among doctors in Bangladesh, which may be due to a lack of attention given to specific food and nutrition topics in their curricula. Additionally, this problem is made worse by the dearth of postgraduate programs that provide doctors and nutritionists with comprehensive training in therapeutic nutrition. While some dietitians do receive education at medical facilities and online, they still don’t have a thorough knowledge of therapeutic diets like the KD. The findings of this study emphasize the need for ongoing training sessions to keep nutritionists up to date on the most recent developments in nutrition science. Before recommending KD to their patients, nutritionists and doctors are strongly advised to take the specialist training on KD offered by the American Nutrition Association (ANA). To improve and refresh their knowledge, we advise doctors and nutritionists to regularly participate in training that is centered on therapeutic nutrition.

Significant correlations between respondents’ knowledge and variables including gender, BMI, education, health profession, and socioeconomic level were found in both univariate and multivariate analyses. However, only gender and BMI showed significant relationships among individuals who had ever used the Ketogenic Diet (Table 3). This is explained by the fact that girls frequently place a greater focus on their weight and physical form. About 73 (96.05%) of the 76 KD followers showed inadequate knowledge levels (see Table 1). Most of these followers had heard of the KD via peers and the internet/social media. They didn’t comprehend the KD’s methodical beginning and ending processes well enough. As a result, the advised methodical strategy of starting the ketogenic diet with a 2:1 ratio of fats and progressively increasing fat consumption was not followed (see Table 5). Referring to Table 4, univariate and multivariate analysis revealed that considerably more respondents who had ever followed the KD thought their weight would stay the same once they stopped doing so. In actuality, though, most people who lose weight through any kind of diet tend to gain it back and occasionally much more than they did before. Only 20% of people who start weight loss attempts while they are already overweight succeed in keeping the weight off in the long run [41]. Due to the ketogenic diet’s extremely restrictive nature, which severely restricts carbohydrate consumption to less than 10% of daily calorie intake, weight rebound after a ketogenic diet is especially common. Following the ketogenic diet can result in weight gain for a variety of causes [41, 42].

Participants in this study were found to have a general lack of understanding, which was consistent with their assumption that the Ketogenic Diet might treat a variety of illnesses (see Table 4). However, there is a contradiction in reality. According to two studies [42, 43], ketogenic diets are not only bad for heart in the long run, but they also make it difficult to lose weight permanently. The study looked at the consequences of keto eating and found a big problem: hardly enough people who start a keto diet stick with it long enough to evaluate its long-term health implications. The inevitable weight rebound therefore negates the short-term benefit of fast weight loss. People who experience this condition acquire more weight in addition to the weight they had previously lost, along with higher cholesterol levels. These studies concede that early weight reduction on the ketogenic diet may momentarily decrease blood pressure and cholesterol levels, which are indications of cardiovascular risk. These indicators do have a tendency to rise again over time, though. This happens as dieters struggle to sustain the weight reduction they first experienced while on the diet. These results are supported by our study, which found that KD followers could only commit to the practice for a maximum of three months (see Figure 2a). This underscores the challenges people encounter maintaining long-term adherence to the KD, which might have a severe impact on their health.

“Keto” refers to ketones, which the body releases when it reaches a state of ketosis, for people who are inexperienced with the diet fads of the previous two decades. By using fat as the body’s major fuel source rather than carbs, this promotes weight reduction more quickly. The ketogenic diet depends on ingesting meals high in fat and protein, frequently taken from animal sources rich in saturated fats. This raises several concerns. These saturated fats are known to raise cholesterol levels, promote the buildup of plaque, which finally results in arterial blockages and may contribute to heart disease. Due to different causes, several respondents gave up the ketogenic diet (as indicated in Figure 2c). The strict restriction on carbohydrate intake typically less than 10% of total calories or even less is one reason why the diet is tough to follow. It’s remarkable that diets like the keto, Paleo, or intermittent fasting, all of which have grown in popularity, may be effectively maintained in laboratory animals. This accomplishment is due to the researchers’ capacity to regulate the animals’ dietary intake. For humans, the circumstance is different. Such diets frequently fail because to real-life occurrences like birthdays, hunger, and the appeal of beloved carb-heavy meals. The best course of action is to embrace a diet high in fruits, vegetables, whole grains, legumes, nuts, and seeds to promote long-term heart health and the pursuit of an active and healthy lifestyle, including the maintenance of a healthy weight. Animal fat consumption must be reduced at the same time.

A thorough review study conducted by Noto et al. in 2013 [44] involving half a million people revealed that a ketogenic diet was associated with higher rates of tumors and hypertension. A ketogenic diet is characterized by restricting carbohydrate intake to less than 50 grams per day, roughly equivalent to the amount found in two apples. According to the study’s findings, low-carb diets were associated with a noticeably greater risk of death from all causes. However, there was no discernible correlation between these diets and the incidence or death of cardiovascular disease (CVD). A ketogenic diet can also have negative impacts on bone and muscular health, as demonstrated in research by Heikura et al. in 2019 and Masood et al. in 2022 [45, 46].

## 5.0 Conclusion

The majority of responders believed that the Keto diet was primarily a way to lose weight. The study’s findings highlight the fact that Bangladeshi people have little understanding of the ketogenic diet’s therapeutic uses and have little awareness of them. Participants showed a lack of knowledge about the possible dangers of the ketogenic diet. To obtain thorough understandings of every aspect and the appropriate methods for putting this diet into practice, it is imperative that everyone receive pre-diet counselling.

## Data Availability

All data produced in the present study are available upon reasonable request to the authors

## Limitations of the Study

Participants’ responses were gathered for the study using an online platform. However, a wider cross-section of the people from the nation could not have been included in this study due to the lack of internet access and practical issues. There were some people whose biases were excluded from the data because of their strict devotion to the Ketogenic Diet. In addition, several participants decided not to finish the research out of fear about possible detrimental effects on their life. In addition, unanticipated events prevented interviews with health experts, which decreased participation from this group. Reaching every part of Bangladesh was difficult due to a lack of financing, which may have prevented a full representation in the study’s findings.

## Authors Contribution

AFT and MMR conceived the study with input from PC and TH. AFT and TH developed questionnaires and study design. AFT led the project regarding volunteer recruitments, data collections to writing with the help of PC and TH. MMR led the analysis of individual-case data and estimation of the onset-to-outcome distributions, with input from AFT, PC and TH. AFT and PC coordinated management of the team, including the data collection, analysis, and processing. AFT, TH and MMR produced the first draft of the manuscript. PC added additional points in discussions. AFT, PC, TH, and MMR finalized the manuscripts after necessary corrections and obtaining suggestions from all authors. MMR supervised all the works from the beginning to the end. All authors did read and agreed unanimously to submit the manuscripts.

## Acknowledgement

The authors would like to thank Mst. Mahfuza Akter from the Department of Community Nutrition at Bangladesh University of Health Sciences in Dhaka, Bangladesh. Her aid in data collecting and insightful contributions during the collaboration efforts are much appreciated.

